# Soluble Immune Checkpoint Protein and Lipid Network Associations with All-Cause Mortality Risk: Trans-Omics for Precision Medicine (TOPMed) Program

**DOI:** 10.1101/2025.01.08.25320225

**Authors:** Annabelle Rodriguez, Chaojie Yang, Wenqi Gan, Keaton Karlinsey, Beiyan Zhou, Stephen S. Rich, Kent D. Taylor, Xiuqing Guo, Jerome I. Rotter, W. Craig Johnson, Elaine Cornell, Russell P. Tracy, J. Peter Durda, Robert E. Gerszten, Clary B. Clish, Thomas Blackwell, George J. Papanicolaou, Honghuang Lin, Laura M. Raffield, Jose D. Vargas, Ramachandran Vasan, Ani Manichaikul

## Abstract

Adverse cardiovascular events are emerging with the use of immune checkpoint therapies in oncology. Using datasets in the Trans-Omics for Precision Medicine program (Multi-Ethnic Study of Atherosclerosis, Jackson Heart Study [JHS], and Framingham Heart Study), we examined the association of immune checkpoint plasma proteins with each other, their associated protein network with high-density lipoprotein cholesterol (HDL-C) and low-density lipoprotein cholesterol (LDL-C), and the association of HDL-C- and LDL-C-associated protein networks with all-cause mortality risk. Plasma levels of LAG3 and HAVCR2 showed statistically significant associations with mortality risk. Colocalization analysis using genome wide-association studies of HDL-C or LDL-C and protein quantitative trait loci from JHS and the Atherosclerosis Risk in Communities identified TFF3 rs60467699 and CD36 rs3211938 variants as significantly colocalized with HDL-C; in contrast, none colocalized with LDL-C. The measurement of plasma LAG3, HAVCR2, and associated proteins plus targeted genotyping may identify patients at increased mortality risk.

---

The approval of relatlimab (a lymphocyte activation gene-3 [LAG3] blocking monoclonal antibody) by the United States Federal Drug Administration (FDA; March 18, 2022) for the indication of metastatic melanoma supports the continued interest and medical need for immunotherapies^1^. A major component of this approval was based on results from the RELATIVITY-047 study, in which investigators reported that the combination of relatlimab and nivolumab (a PD-1 blocking monoclonal antibody) significantly improved progression-free survival as compared with nivolumab alone^2^. The side effect profile of Rel/Niv was as expected, as was the number of immune-mediated adverse events.

One category of adverse events that was not reported in this study was the rate of myocardial infarction (MI). This is of importance given that we have previously reported in the Multi-Ethnic Study of Atherosclerosis (MESA) that LAG3 deficiency is not rare and is an independent predictor of high-density lipoprotein cholesterol (HDL-C) and increased cardiovascular disease (CVD) risk^3^. In subjects with HDL-C ≥60 mg/dl, low plasma LAG3 protein levels significantly increased MI risk and plasma LAG3 added predictive value to the Framingham risk score. This relationship of high HDL-C with increased CVD risk is considered paradoxical and multiple studies have reported this observation^4–7^. In the Reasons for Geographic and Racial Differences in Stroke (REGARDS) study, investigators reported that high HDL-C levels were not protective against CVD risk in either Black or white subjects^8^. The underlying etiologies for this high HDL-C paradoxical phenotype have not been thoroughly identified.

An observational study from a tertiary academic center sought to determine newly diagnosed CVD cases in patients treated with immune checkpoint inhibitors (ICIs) for cancer treatment^9^. Data from an electronic medical record system were used to determine the rates of CVD in patients treated with FDA approved immunotherapies (e.g., anti-CTLA-4, anti-PD-1) and/or anti-PDCD1LG1. Out of 102,701 patients, 424 patients were treated with at least one ICI and, of these patients, 62 (14.6%) were newly diagnosed with CVD. In these newly diagnosed cases, the most frequent conditions were arrhythmia and heart failure, with single use of anti-CTLA-4 (compared with the other ICIs) correlated with higher rates of heart failure. In addition, those patients with a history of ischemic heart disease had a higher risk of mortality during the course of cancer treatment.

The objective of this study was to leverage the datasets available in the Trans-Omics for Precision Medicine (TOPMed) program to determine the association of available soluble immune checkpoint proteins (LAG3, CTLA-4 [cytotoxic T-lymphocyte associated protein 4], PDCD1LG1 [programmed cell death protein 1 ligand 1], PDCD1LG2 [programmed cell death protein 1 ligand 2], CD40LG [CD40 ligand], and HAVCR2 [hepatis A virus cellular receptor 2])and their associated protein networks with major lipid classes and all-cause mortality risk. The analyses included plasma proteomics from MESA (a multi-ethnic population), the Jackson Heart Study (JHS, an African American population) and the Framingham Heart Study (FHS, a mostly white population). Additionally, we performed a meta-analysis of protein quantitative trail loci (pQTLs) from JHS and the Atherosclerosis Risk in Communities (ARIC) study^10^ followed by genetic colocalization analysis to identify overlapping variants for proteins and HDL-C or LDL-C.

## METHODS

### Trans-Omics for Precision Medicine (TOPMed) program

TOPMed is sponsored by the National Heart Lung Blood Institute to harmonize across multiple large, well-phenotyped cohorts related to cardiopulmonary disease with deep WGS and other –omics datasets (proteomics [SomaScan^R^], among others) (https://topmed.nhlbi.nih.gov/). Access for datasets generated by TOPMed for MESA, JHS, and FHS has dbGaP approval (project #21279). The rationale for examining these three cohorts was based on self-described race/ethnic diversity and availability of the -omics, outcome, and demographic data.

### Study populations

The Multi-Ethnic Study of Atherosclerosis (MESA) is an ongoing longitudinal study of the natural history of atherosclerotic disease in multi-ethnic populations from six sites in the US^11–13^. The self-reported representative populations analyzed in this report include Whites, African Americans, Hispanic/Latino, and Asian (predominantly Chinese American) (**Table 1**). The Jackson Heart Study (JHS) is a longitudinal community-based cohort study begun in 2000 consisting of self-described African American individuals from Jackson, MS^14^. The Framingham Heart Study (FHS) is a community-based cohort initiated in 1948. Three generations of participants have been recruited, and the majority of them were self-described white individuals of European ancestry. Participants were invited to attend physical examinations every 2–8 years. The current study was restricted to the Offspring cohort participants who attended their fifth clinical examination cycle during 1991–1995^15^.

**Table 1A.** Study population demographics for MESA, JHS, and FHS:Offspring.

| **Characteristics** | **MESA** | **JHS** | **FHS:Offspring** |
| --- | --- | --- | --- |
| **No. subjects** | 6429 | 5306 | 3821 |
| **Women (N, %)** | 3369 (52) | 3367 (63.4) | 1927 (50.4) |
| **Race/Ethnicity (N, %)** |  |  |  |
| **European White** | 2527 (39) | na | 3821 (100) |
| **African-American** | 1677 (26) | 5306 (100) | na |
| **Hispanic** | 1450 (23) | na | na |
| **Asian-American** | 775 (12) | na | na |
| **Age, years (mean ± STD)** | 62.2 ± 10.2 | 54.8 ± 12.9 | 53.8 ± 9.5 |
| **Total Cholesterol, mg/dl** | 194.4 ± 35.8 | 199.3 ± 40.1 | 203.9 ± 36.5 |
| **HDL-C, mg/dl** | 50.9 ± 14.8 | 51.8 ± 14.6 | 50.2 ± 15.1 |
| **LDL-C, mg/dl** | 117.3 ± 31.5 | 126.6 ± 36.6 | 125.8 ± 32.6 |
| **Triglycerides, mg/dl** | 132.6 ± 89.7 | 106.4 ± 78.4 | 145.3 ± 116.0 |
| **BMI, kg/m<sup>2</sup>** | 28.3 ± 5.5 | 31.8 ± 7.2 | 27.3 ± 4.9 |
| **Fasting glucose, mg/dl** | 97.4 ± 30.5 | 100.0 ± 33.4 | 99.6 ± 25.2 |
| **Serum creatinine, mg/dl** | 0.96 ± 0.29 | 1.04 ± 0.52 | 1.06 ± 0.27 |
| **Systolic BP, mmHg** | 134.2 ± 19.0 | 127.5 ± 16.9 | 125.1 ± 18.6 |
| **Diastolic BP, mmHg** | 75.2 ± 10.1 | 75.7 ± 8.8 | 74.5 ± 10.1 |
| **Cigs per day or smoking history** | 16.3 ± 17.5 | No (87%) | 3.74 ± 9.4 |
| **Alcohol use or history** | 0.69 ± 0.46 | No (54%) | 1.02 ± 1.21 |
| **All-cause mortality (N, %)** | 1612 (25) | 1378 (26) | 1627 (42.6) |
| **Protein targets**<br>**(SomaScan), RFU** | N=1276 | N=1322 | N=1314 |
| **Number of subjects with**<br>**SomaScan measurements** | N=934 | N=2143 | N=851 |

Abbreviations: HDL-C= high-density lipoprotein cholesterol; LDL-C= low-density lipoprotein cholesterol; BMI= body mass index; BP=blood pressure; Alcohol use= number of drinks per week; RFU=relative fluorescent units; NA=not applicable

All MESA participants provided written informed consent for participation at the six field sites, JHS participants provided written informed consent for participation at the single site in Jackson, MS, and FHS participants provided written informed consent for participation at the single site in Framingham, MA. MESA, JHS, and FHS study protocols were reviewed and approved by the Institutional Review Boards at each of the participating study sites, with this specific analysis also approved at the University of Connecticut Health, Farmington, CT (AR).

### Genetic and phenotypic data

TOPMed phenotypic datasets were available across each of the cohorts as reported in the TOPMed website. Whole genome sequencing (WGS) was performed at multiple sequencing centers (Broad Institute, University of Washington) at a mean depth of >30X using Illumina HiSeq X Ten instruments, with variant quality control performed by the TOPMed Informatics Research Center (IRC, University of Michigan) using the GotCloud pipeline^16^. SomaScan proteomics used DNA aptamers that bind to protein epitopes and was quantified by hybridization to DNA microarrays, with units reported as relative fluorescent units (RFU)^16–19^.

Phenotypic data from MESA was obtained from Exam 1 (2000-2002) and Exam 2 (2002-2004). SomaScan proteomics was available for 934 subjects (n=1276 protein targets) at Exam 1^20^ while plasma LAG3 was also measured by ELISA in 5622 subjects from Exam 2^3, 17, 21^. Clinical outcomes (e.g., all-cause mortality and relevant covariates) were obtained through 2018 and were adjudicated as previously described^12, 13^. In JHS, SomaScan proteomics was available for 2143 subjects (n=1322 protein targets)^19^ at Exam 1 (across three batches) and survival data was updated through 2018. In FHS, SomaScan proteomics (n=1314 proteins, batch 2) was available for 851 subjects from the Framingham Offspring Cohort Exam 5 (1991-1995)^22^ and survival data was updated through 2018.

### Regression analyses

A linear regression analysis was performed with each of the individual six immune checkpoint proteins (LAG3, CTLA-4, PDCD1LG1, PDCD1LG2, HAVCR2, CD40LG) as the outcome in models adjusted for age and sex (**Table 1B**). All proteins were transformed using ranked inverse normal transformation and then entered separately into the regression models. The proteomics data for JHS was further adjusted for batch effects. For association studies examining the independent effect of the proteins on lipids (total cholesterol, LDL-C, HDL-C, and triglycerides), models were adjusted for age, sex, race/ethnicity, and principal components of ancestry (PCA) [only in MESA], lipids [excluded if lipid was the outcome], body mass index (BMI), smoking, alcohol consumption, serum creatinine, fasting blood glucose, and lipid medication use. For association studies examining the independent effect of the proteins on all-cause mortality, models were adjusted for age, sex, race/ethnicity and principal components of ancestry (PCA) [only in MESA], total cholesterol, BMI, hypertension, smoking, alcohol consumption, serum creatinine, fasting blood glucose, systolic blood pressure, and diastolic blood pressure. Individual proteins were entered separately into the models, with and without an interaction term for LAG3, PDCD1LG1, PDCD1LG2 or HAVCR2. In each cohort, for the lipid analyses, linear regressions were first performed. Benjamini-Hochberg false discovery rate (FDR) was applied to all proteins and then those proteins with adjusted *P* value ≤0.05 were used in the all-cause mortality association analyses. As an example, in MESA, of the 1275 proteins associated with LAG3, 845 were significant based on FDR ≤0.05. Of these 845 LAG3 proteins, 35 were significantly associated with HDL-C, and none were significantly associated with all-cause mortality. This same strategy was used for the other cohorts. We also uploaded the list of proteins associated with mortality into the STRING database and performed a cluster analysis to identify the association of these proteins with known protein-protein interaction networks in STRING^23^.

**Table 1B.**
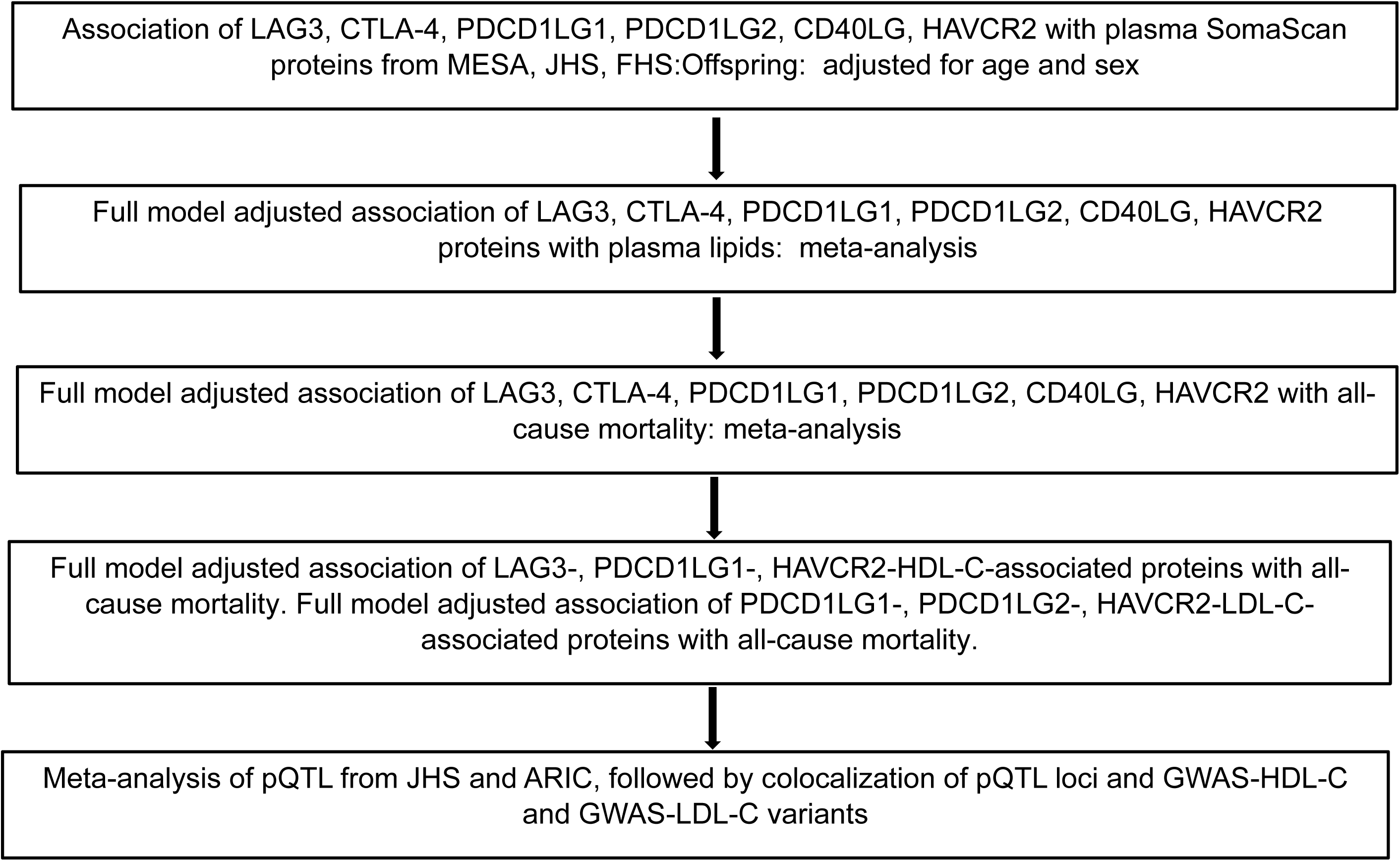
Analysis Workflow Schematic.

### Bayesian colocalization analyses

Meta-analysis of pQTLs from JHS^24^ and ARIC^10^ in African Americans was performed using METAL^25^. Colocalization analysis was performed using the “coloc” R package^26,27^, leveraging pQTL and GWAS of HDL-C^28^ and LDL-C^28^ to identify the variants colocalizing with HDL-C and LDL-C and proteins. Colocalization was based on a genome window size of ±1 Mb of the transcriptional start sites for each protein (LAG3-HDL-C-, PDCD1LG1-HDL-C-, PDCD1LG1-LDL-C, PDCD1LG2-LDL-C- and HAVCR2-HDL-C-, and HAVCR2-LDL-C-mortality-associated proteins). Bayesian colocalization analysis tested four hypotheses - H0: neither GWAS nor pQTL has a genetic association in the region; H1: only GWAS has a genetic association in the region; H2: only pQTL has a genetic association in the region; H3. both GWAS and pQTL are associated, but with different causal variants; and H4: both GWAS and pQTL are associated and share a single causal variant. A posterior colocalization probability of H4 greater than 0.80 (PP.H4 > 0.80) was used as the threshold for colocalization visualization.

### Statistical analyses

Triglyceride levels were log-transformed prior to inclusion in regression analyses. False discover rate (FDR) ≤ 0.05 was used as the threshold for statistical significance. JMP v17.0, and R were the statistical software packages used for these analyses. Meta-analyses (the *‘metafor’* R package^29^) were used to synthesize the original results from individual cohorts. Heterogeneity of results across studies used a Cochran Q test. If significant heterogeneity was present (*P* < 0.10), a random-effects model was used; in the absence of significant heterogeneity, a fixed-effects model was used.

### Data availability

TOPMed multi-omics datasets are available following approval of a TOPMed paper proposal and an approved dbGaP (database of genotypes and phenotypes) project. dbGaP accession numbers are phs000209.v13.p3 and phs001416.v3.p1 (MESA), phs000286.v6.p2 and phs000964.v5.p1 (JHS), and phs000007.v33.p1 and phs000974.v5.p4 (FHS).

## RESULTS

### Plasma immune checkpoint protein-associated networks

GeneAnalytics^TM^, a gene set enrichment statistical analysis tool, was used to identify the rank and score of genes corresponding to all SomaScan proteins in each cohort^30^. For all three cohorts, the highest matched pathways were innate immune system and signal transduction (*P*≤0.0001) (**Supplementary Tables SI-SIII**).

In all three cohorts, six soluble immune checkpoint proteins (LAG3, CTLA-4, PDCD1LG1, PDCD1LG2, CD40LG, and HAVCR2 but not PDCD1, TIGIT or BTLA) were identified in the SomaScan proteomics datasets. The association of these six immune checkpoint proteins with each other were examined in a model adjusted for age, sex and PCA (MESA only). Meta-analysis revealed that plasma CTLA-4 was significantly associated with PDCD1LG1 (*P*=0.018) and HAVCR2 (*P*=0.019) but not with the other immune checkpoint proteins, while the rest were significantly associated with each other (**Table 2**).

**Table 2.** Associations of soluble immune checkpoint proteins with each other: meta-analysis of MESA, JHS, and FHS Offspring.

| **Protein** | **$\beta$ (95% CI)** | ***P* Value** |
| --- | --- | --- |
| **LAG3** |  |  |
| CTLA-4 | 0.07 (-0.01 to 0.15) | 0.081 |
| PDCD1LG1 | 0.16 (0.05 to 0.27) | **0.004** |
| PDCD1LG2 | 0.16 (0.09 to 0.23) | **< 0.001** |
| CD40LG | -0.12 (-0.15 to -0.09) | **< 0.001** |
| HAVCR2 | 0.30 (0.21 to 0.39) | **< 0.001** |
| **CTLA-4** |  |  |
| LAG3 | 0.08 (-0.01 to 0.16) | 0.088 |
| PDCD1LG1 | 0.10 (0.02 to 0.19) | **0.018** |
| PDCD1LG2 | -0.03 (-0.05 to 0.00) | 0.069 |
| CD40LG | -0.08 (-0.23 to 0.07) | 0.276 |
| HAVCR2 | 0.09 (0.01 to 0.16) | **0.019** |
| **PDCD1LG1** |  |  |
| LAG3 | 0.17 (0.06 to 0.28) | **0.003** |
| CTLA-4 | 0.10 (0.02 to 0.19) | **0.018** |
| PDCD1LG2 | 0.10 (0.01 to 0.19) | **0.036** |
| CD40LG | -0.16 (-0.23 to -0.10) | **< 0.001** |
| HAVCR2 | 0.25 (0.15 to 0.35) | **< 0.001** |
| **PDCD1LG2** |  |  |
| LAG3 | 0.17 (0.09 to 0.24) | **< 0.001** |
| CTLA-4 | -0.03 (-0.05 to 0.00) | 0.069 |
| PDCD1LG1 | 0.10 (0.01 to 0.19) | **0.036** |
| CD40LG | -0.10 (-0.13 to -0.07) | **< 0.001** |
| HAVCR2 | 0.32 (0.29 to 0.35) | **< 0.001** |
| **CD40LG** |  |  |
| LAG3 | -0.13 (-0.16 to -0.10) | **< 0.001** |
| CTLA-4 | -0.08 (-0.23 to 0.07) | 0.276 |
| PDCD1LG1 | -0.16 (-0.24 to -0.09) | **< 0.001** |
| PDCD1LG2 | -0.10 (-0.13 to -0.07) | **< 0.001** |
| HAVCR2 | -0.20 (-0.26 to -0.13) | **< 0.001** |
| **HAVCR2** |  |  |
| LAG3 | 0.29 (0.21 to 0.37) | **< 0.001** |
| CTLA-4 | 0.07 (0.01 to 0.14) | **0.017** |
| PDCD1LG1 | 0.22 (0.13 to 0.31) | **< 0.001** |
| PDCD1LG2 | 0.29 (0.26 to 0.31) | **< 0.001** |
| CD40LG | -0.18 (-0.24 to -0.11) | **< 0.001** |

### Association of plasma immune checkpoint proteins with lipids

Having previously identified an inverse association of LAG3 with HDL-C in MESA participants, we performed a meta-analysis on LAG3, CTLA-4, PDCD1LG1, PDCD1LG2, CD40LG and HAVCR2 with total cholesterol, HDL-C, LDL-C, and triglycerides across the three cohorts (the lipid association analyses for each cohort are shown in **Supplementary Table SIV**). LAG3 was significantly associated with total cholesterol (beta −1.74, *P*=6.47E-03), HDL-C (beta −2.10, *P=*4.54E-23) and triglycerides (beta −0.03, *P*=3.3E-02), PDCD1LG1 was significantly associated with all lipid groups (total cholesterol: beta −3.17, *P=*3.36E-07; HDL-C: beta −0.55, *P*=1.01E-02; LDL-C: −2.69, *P*=5.39E-06; triglycerides: −0.02, *P*=1.3E-02), PDCD1LG2 was significantly associated with total cholesterol (beta −1.86, *P*=3.70E-03), LDL-C (beta −1.55, *P*=1.08E-02) and triglycerides (beta −0.03, *P*=1.1E-04), HAVCR2 was significantly associated with all lipid groups (total cholesterol: beta −5.15, *P=*7.35E-15; HDL-C: beta −0.81, *P*=3.24E-02; LDL-C: −4.60, *P*=2.81E-13; triglycerides: −0.03, *P*=4.5E-06), while CTLA-4 and CD40LG were not significantly associated with any of the major lipid groups (**Table 3**).

**Table 3.**
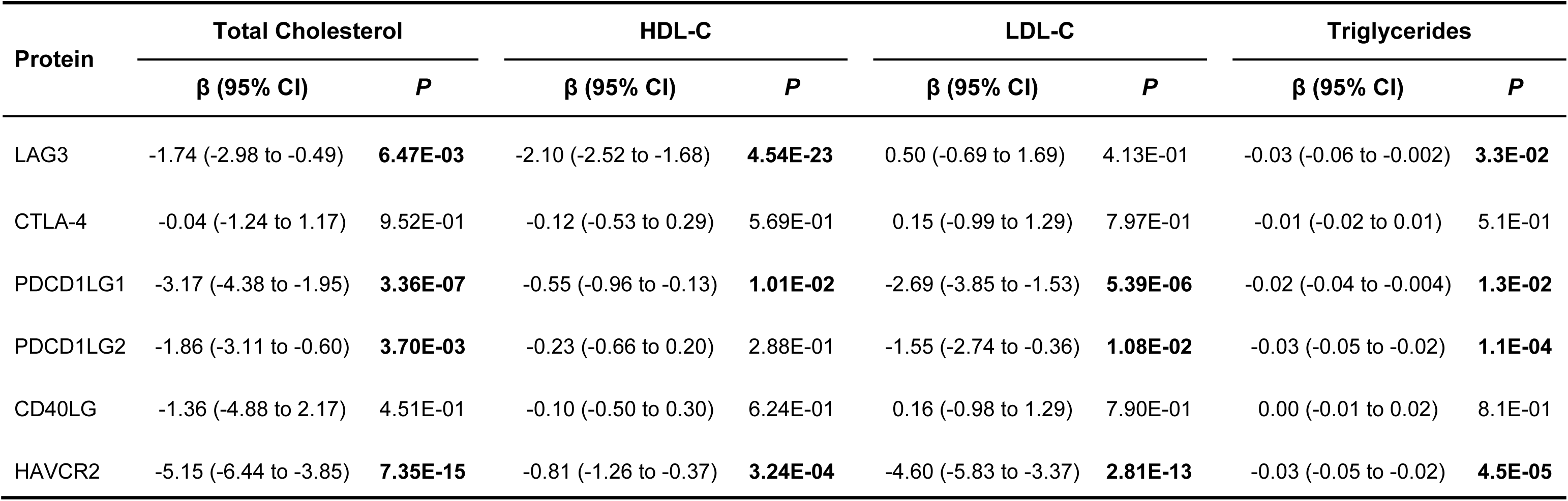
Associations of immune checkpoint proteins with major lipid classes: meta-analysis of MESA, JHS, and FHS Offspring.

| Protein | Total Cholesterol |  | HDL-C |  | LDL-C |  | Triglycerides |  |
| --- | --- | --- | --- | --- | --- | --- | --- | --- |
| | $\beta$ (95% CI) | *P* | $\beta$ (95% CI) | *P* | $\beta$ (95% CI) | *P* | $\beta$ (95% CI) | *P* |
| LAG3 | -1.74 (-2.98 to -0.49) | **6.47E-03** | -2.10 (-2.52 to -1.68) | **4.54E-23** | 0.50 (-0.69 to 1.69) | 4.13E-01 | -0.03 (-0.06 to -0.002) | **3.3E-02** |
| CTLA-4 | -0.04 (-1.24 to 1.17) | 9.52E-01 | -0.12 (-0.53 to 0.29) | 5.69E-01 | 0.15 (-0.99 to 1.29) | 7.97E-01 | -0.01 (-0.02 to 0.01) | 5.1E-01 |
| PDCD1LG1 | -3.17 (-4.38 to -1.95) | **3.36E-07** | -0.55 (-0.96 to -0.13) | **1.01E-02** | -2.69 (-3.85 to -1.53) | **5.39E-06** | -0.02 (-0.04 to -0.004) | **1.3E-02** |
| PDCD1LG2 | -1.86 (-3.11 to -0.60) | **3.70E-03** | -0.23 (-0.66 to 0.20) | 2.88E-01 | -1.55 (-2.74 to -0.36) | **1.08E-02** | -0.03 (-0.05 to -0.02) | **1.1E-04** |
| CD40LG | -1.36 (-4.88 to 2.17) | 4.51E-01 | -0.10 (-0.50 to 0.30) | 6.24E-01 | 0.16 (-0.98 to 1.29) | 7.90E-01 | 0.00 (-0.01 to 0.02) | 8.1E-01 |
| HAVCR2 | -5.15 (-6.44 to -3.85) | **7.35E-15** | -0.81 (-1.26 to -0.37) | **3.24E-04** | -4.60 (-5.83 to -3.37) | **2.81E-13** | -0.03 (-0.05 to -0.02) | **4.5E-05** |

The lipid meta-analysis results have validated the inverse association of HDL-C with plasma LAG3 and additionally identified PDCD1LG1 and HAVCR2 as inversely associated with HDL-C. To identify a network of SomaScan plasma proteins significantly associated with LAG3 and HDL-C, for each cohort, we first performed a regression analysis of LAG3 as the dependent variable in a model including separate inclusion of SomaScan proteins and corrected for FDR≤0.05 (**Supplementary Table SV-VII, Table 1B**). A second regression analysis was performed with HDL-C as the dependent variable in a fully adjusted model with inclusion of the LAG3-associated proteins (each entered separately, **Supplementary Table SVIII-SX**). Meta-analysis for LAG3-HDL-C associated proteins from the three cohorts identified thirty-two proteins that were significantly associated with HDL-C (**Supplementary Table SXI**), with seventeen negatively and fifteen positively associated with HDL-C (meta-analyses of PDCD1LG1-HDL-C and HAVCR2-HDL-C-associated proteins are found in **Supplementary Tables SXII-SXXIV**).

### Effect of immune checkpoint proteins on all-cause mortality risk

The association of each of the immune checkpoint proteins with all-cause mortality was interrogated in each cohort using a Cox proportional hazard analysis model with all-cause mortality as the outcome with each transformed protein (LAG3, CTLA-4, PDCD1LG1, PDCD1LG2, CD40LG, HAVCR2) entered separately. Meta analysis identified HAVCR2 as significantly associated with mortality with hazard ratio [HR] 1.21 (confidence interval [CI]: lower 95% 1.12, upper 95% 1.32, *P* =4.85×10^-6^) (**Table 4).** Results from the Cox proportional hazard analyses of each cohort are shown in **Supplementary Table SXXV**.

**Table 4.** All-cause mortality of immune checkpoint proteins: meta-analysis of MESA, JHS, and FHS Offspring.

|  | **Hazard Ratio** | **Lower 95% CI** | **Upper 95% CI** | **P Value** |
| --- | --- | --- | --- | --- |
| LAG3 | 0.96 | 0.89 | 1.03 | 2.70E-01 |
| CTLA-4 | 1.00 | 0.93 | 1.07 | 8.90E-01 |
| PDCD1LG1 | 1.05 | 0.98 | 1.13 | 1.80E-01 |
| PDCD1LG2 | 1.06 | 0.87 | 1.31 | 5.50E-01 |
| CD40LG | 1.02 | 0.95 | 1.10 | 5.50E-01 |
| HAVCR2 | 1.21 | 1.12 | 1.32 | **4.85E-06** |

Since the paradox of high HDL-C and increased all-cause mortality risk has been reported in large observational studies and there is literature supporting significant differences in HDL-C based on self-reported race/ethnicity^31–33^, Cox proportional hazard analyses were performed using the same fully adjusted model including plasma proteins that were identified as significantly associated with LAG3 and HDL-C. Each protein was separately entered in the model containing LAG3-HDL-C associated protein and its corresponding interaction term, and this analysis was done separately for each cohort. In MESA, the results showed that none of the thirty-five proteins were significantly associated with all-cause mortality based on FDR≤0.05. In MESA, in addition to SomaScan plasma LAG3 measurements, we also had availability of plasma LAG3 as measured by an ELISA in a larger subset of the cohort at Exam 2 (n=5622 total, n=1442 in subjects with HDL-C ≥60 mg/dl)^3^. A hazard analysis found that plasma LAG3 was inversely associated with all-cause mortality risk in subjects with HDL-C ≥60 mg/dl (HR 0.82 [0.68-0.99 lower 95%-upper 95% CI], *P*=0.036, n=303 events and n=999 censorings) (**Figure 1**).

**Figure 1.**
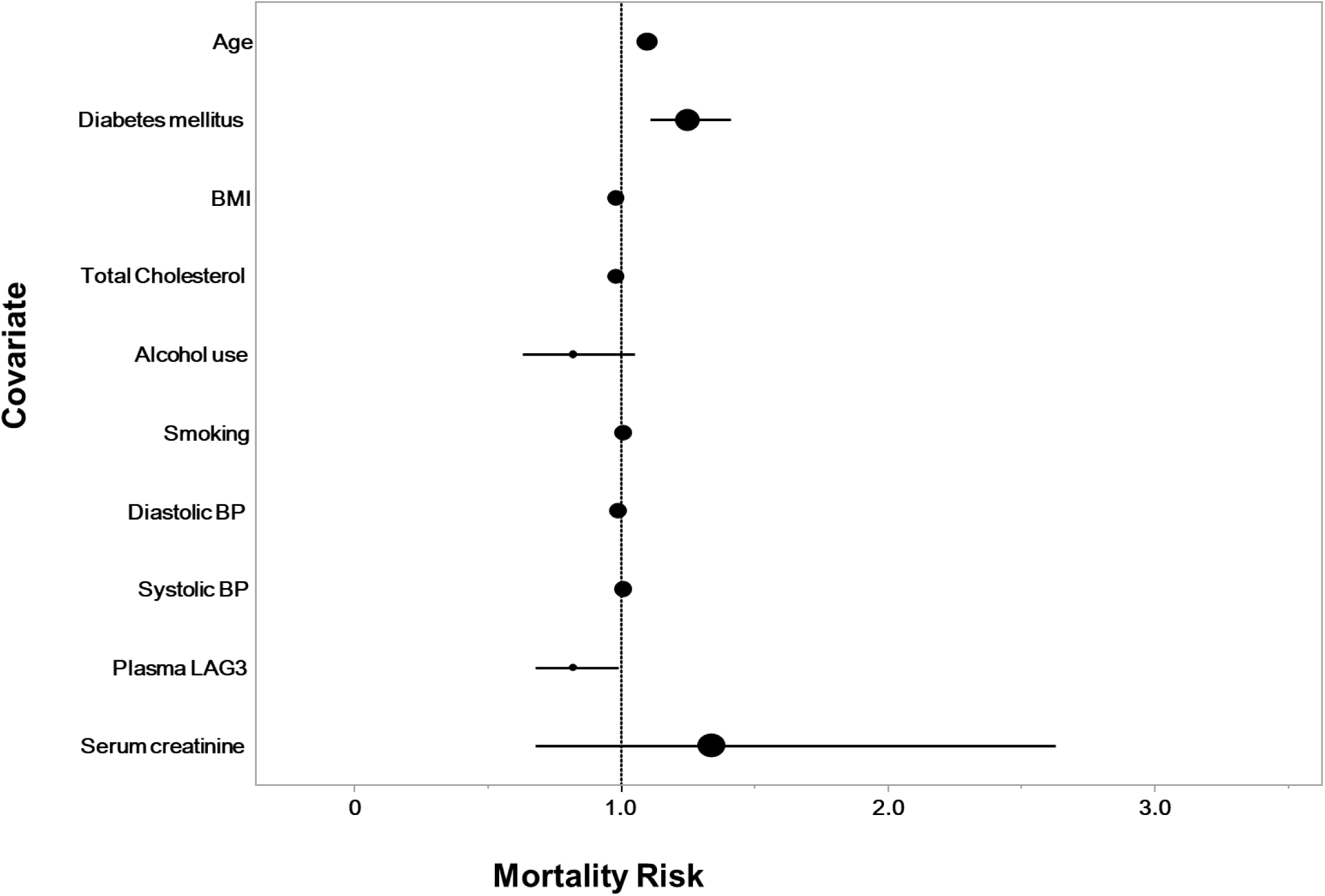
Forest plot of plasma LAG3 significantly associated with mortality risk in MESA subset stratified by HDL-C ≥60 mg/dl. Forest plot of mortality risk of plasma LAG3 as measured by ELISA in a fully adjusted model (HR 0.82 [0.68-0.99 lower 95%-upper 95% CI], *P*=0.036, n=303 events and n=999 censorings)). This was analyzed in MESA subjects stratified by HDL-C ≥60 mg/dl (N=1442).

### LAG3-HDL-C-associated proteins and all-cause mortality risk

In MESA, there were no LAG3-HDL-C-associated proteins associated with mortality risk (n=101 events and n=610 censorings), while in FHS, we identified one protein (PARC: HR 1.34 [1.14-1.56 CI], *P*=0.0003, n=188 events, n=507 censorings) that was significantly associated with mortality risk. In JHS, we identified thirty-seven proteins (including PARC) associated with mortality risk (n=510 events and 1429 censorings) (**Supplementary Table SXXVI)**. PARC exhibits a phenotype of low HDL-C-high mortality risk.

### HAVCR2-HDL-C and HAVCR2-LDL-C-associated proteins and all-cause mortality risk

Our novel observation that HAVCR2 alone was significantly associated with both HDL-C and mortality risk prompted a similar analysis to determine if any of the HAVCR2-HDL-C-associated proteins were significantly associated with mortality risk. None of the HAVCR2-HDL-C-associated proteins in either MESA or FHS were significantly associated with mortality risk; however, forty-four proteins were significantly associated with mortality risk in JHS (**Supplementary Table SXXVII**). We conducted similar analyses of HAVCR2-LDL-C associated proteins and observed that MIC1 alone (HR 1.69 [1.39-2.05, lower-upper 95% CI], *P*<0.0001) was significantly associated with mortality risk in FHS.

### PDCD1LG1-HDL-C associated proteins and all-cause mortality risk

Given that we have now identified PDCD1LG1 as significantly associated with HDL-C (**Table 2**) and the expression of this PDCD1 ligand was measured in the RELATIVITY trial^2^, we conducted identical analyses but now focused on PDCD1LG1-HDL-C associated proteins. In MESA and FHS a survival analysis did not identify any PDCD1LG1-HDL-C associated proteins as significantly associated with mortality risk using FDR≤0.05. However, in JHS, we identified thirty-seven proteins as being significantly associated with mortality risk (**Supplementary Table SXXVIII)**.

### All HDL-C-associated mortality proteins

In JHS, across the LAG3, PDCD1LG1, and HAVCR2 groupings, sixty-one HDL-C-associated proteins were independently associated with mortality risk. The sixty-one proteins were categorized into four phenotypic groups based on their respective HDL-C beta estimate and hazard ratio: low HDL-C_high mortality; high HDL- C_low mortality; low HDL-C_low mortality; and high HDL-C_high mortality (**Supplementary Table SXXIX**). Six proteins exhibited the paradoxical high HDL-C_high mortality risk phenotype (REG4, IGFBP1, MIP5, MIC1, TFF3, IGFBP2). Nineteen exhibited the “protective” high HDL- C_low mortality risk phenotype (INTEGRIN AVB5, CD36, COAGULATION FACTOR XA, WFKN2, CARBONIC ANHYDRASE 6, ANGIOTENSINOGEN, THROMBIN, NCAM120, PMPR1A, ERBB3, TRKC, ERBB1, PROTEIN C, ASAH2, RET, KALLISTATIN, PROTHROMBIN, CNDP1). Twenty-two exhibited the low HDL-C_high mortality risk (IGFBP7, NET4, IGA, C9, CATHEPSIN S, LGMN, ALPHA 1 ANTICHYMOTRYPSIN, MMP12, ANGIOPOETIN 2, CD30, TNFSRI, TNFSF15, MIP1A, PARC, TNFSRII, TSP2, BLC, PROTEINASE 3, DDIMER, HAVCR2, DELL1, LEG9). Fourteen exhibited the low HDL-C_low mortality risk (MYOSTATIN, ATS13, CD200R1, NOGO RECEPTOR, ROBO2, IFNGF1, SOMATOSTATIN28, LRP1B, GDF11_8, A2 ANTIPLASMIN, MYOGLOBIN, FACTOR H, SEMAPHOREIN 6A, MP2K4).

### PDCD1LG1-LDL-C-associated and PDCD1LG2-LDL-C-associated proteins and all-cause mortality risk

Having observed that PDCD1LG1 and PDCD1LG2 were significantly associated with each other (**Table 2**) and LDL-C (**Table 3**), we next examined their respective protein associations and mortality risk. For PDCD1LG1, in MESA, TNFSF15 alone (HR 1.76 [1.36-2.29, lower-upper 95%]) was significantly associated with mortality risk. No PDCD1LG2-LDL-C-associated proteins were associated with mortality risk. In JHS, we identified forty-two PDCD1LG1-LDL-C-associated proteins as significantly associated with mortality risk (**Supplementary Table SXXX**). For PDCD1LG2, we identified thirty-seven proteins as significantly associated with mortality risk (**Supplementary Table SXXXI**). In FHS, for PDCD1LG1-LDL-C-associated proteins, MIC1 was the only protein significantly associated with mortality risk (HR 1.72 [1.41-2.08, lower-upper 95% CI], *P*<0.0001). For PDCD1LG2-LDL-C-associated proteins, none were significantly associated with mortality risk.

### All LDL-C-associated mortality proteins

In JHS, across the PDCD1LG1 and PDCD1LG2 groupings, fifty-two LDL-C-associated proteins were independently associated with mortality risk (including MIC1). The fifty-two proteins were categorized into four phenotypic groups based on their respective LDL-C beta estimate and hazard ratio: low LDL-C_high mortality; high LDL- C_low mortality; low LDL-C_low mortality; and high LDL-C_high mortality (**Supplementary Table SXXXII**). We identified one protein, NET4, that exhibited a high LDL-C_high mortality risk phenotype. Eighteen proteins exhibited a high LDL-C_low mortality risk phenotype (SCFSR, ERBB3, CKMM, ERBB1, PROTEIN C, A2 ANTIPLASMIN, DKK4, SEPR, PROTHROMBIN, FGF20, FGF9, IL1RRP2, CNTN2, CARBONIC ANHYDRASE 6, PGCB, NCAM120, TRKB, CONTACTIN5). Thirty-two proteins exhibited a low LDL-C_high mortality risk phenotype (IGA, C7, CATHEPSIN B, TSP2, PROTEINASE 3, SECTM1, PSP, LTBP4, UCRP, IMP1, ANGIOPOETIN 2, CD30, TNFSRI, LIPOCALIN2, TNSFSF15, RESISTIN, TNFSRII, FCG3B, FSTL3, B2 MICROGLOBULIN, MIP5, A1 ANTITRYPSIN, MIC1, TFF3, CATZ, CLM6, TIMD3, CALGRANULIN B, DLL1, N TERMINAL PROBNP, IGFBP2, FBLN3). Only one protein, RET, exhibited a low LDL-C_low mortality risk phenotype.

Of these fifty-two proteins, twenty-three matched with the HDL-C mortality proteins including IGA, ERBB3, ERBB1, PROTEIN C, A2 ANTIPLASMIN, TSP2, PROTEINASE 3, PROTHROMBIN, ANGIOPOIETIN 2, CD30, TNFSRI, TNFSF15, TNFSRII, RET, NET4, CARBONIC ANHYDRASE 6, MIP5, MIC1, NCAM120, TFF3, HAVCR2, DLL1, IGFBP2 (STRING protein-protein cluster interaction map, **Supplementary Figure S1**). The unique LDL-C mortality associated proteins were CSF2RA, CKMM, C7, CATHEPSIN B, CNTN5, DKK4, SEPR, SECTM1, PSP, LTBP4, UCRP, TIMP1, FGF20, FGF9, LCN2, IL1RRP2, RESISTIN, CNTN2, FCGR3B, FSTL3, BCAN, B2 MICROGLOBULIN, SERPINA1, TRKB, CAT, CD300C, CALGRANULIN B, NPPB, FBLN3 (**Supplementary Figure S2**). The unique HDL-C mortality associated proteins were MYOSTATIN, IGFBP7, INTEGRIN AVB5, CD36, C9, COAGULATION FACTOR XA, ATS13, CATHEPSIN S, WFKN2, ANGIOTENSINOGEN, LGMN, ALPHA 1 ANTICHYMOTRYPSIN, THROMBIN, MMP12, BMPR1A, CD200R1, NOGO RECEPTOR, ROBO2, IFNGF1, SOMATOSTATIN28, LRP1B, REG4, TRKC, GDF11_8, IGFBP1, MIP1A, MYOGLOBIN, PARC, ASAH2, KALLISTATIN, BLC, FACTOR H, KIRR3, DDIMER, SEMAPHORIN 6A, MP2K4, CNDP1, LEG9 (**Supplementary Figure S3**).

### Quantitative trait loci and colocalization analyses of proteins associated with LAG3, PDCD1LG1, PDCD1LG2, HAVCR2, lipids and mortality risk in JHS

Thus far the largest number of candidate proteins associated with lipids and mortality risk have been found in the JHS population, in part due to the larger sample size in JHS compared with MESA and FHS. For the lipid associated mortality risk proteins, we first performed meta-analysis of pQTLs from JHS^19^ and ARIC^10^ and then performed colocalization analysis using this meta-analyzed pQTL and GWAS of HDL-C and LDL-C to identify which colocalized based on the threshold of PPH4 >0.80^27^. Based on these analyses, Trefoil factor 3 (*TFF3* gene, TFF3 protein), one of the mortality risk proteins with the paradoxical high HDL-C_high mortality phenotype, showed evidence of genetic colocalization with HDL-C (PPH4 = 88.8%), with the most significant shared variant (rs60467699) having associations with HDL-C (*P* = 4.44E-13) and pQTL (*P* = 1.46E-11) (**Figure 2, panel A**). The rs60467699 is characterized as A>G, 0.09% frequency, chromosome position 21: 42,301,959, 3’ UTR downstream variant of the *TFF3* locus. The *TFF3* locus is adjacent to *ABCG1* (21: 42,219,140), a gene known to regulate HDL-C levels. Additionally, colocalization analysis also identified that CD36 (scavenger receptor class B type III, *CD36 gene,* CD36 protein), one of the HAVCR2-HDL-C-mortality risk proteins, showed evidence of colocalization with HDL-C (PPH4 = 98.8%), with the most significant missense variant (rs3211938) colocalized with HDL-C (*P* = 4.18E-55) and pQTL (P = 4.75E-221). The association of CD36 with HAVCR2-HDL-C showed a beta estimate of 1.31 and a HR of 0.89, indicating the HDL/mortality phenotype was higher CD36 plasma levels with higher HDL-C levels and decreased mortality risk. We did not observe significant genetic colocalization of the LDL-C-mortality associated proteins.

**Figure 2.**
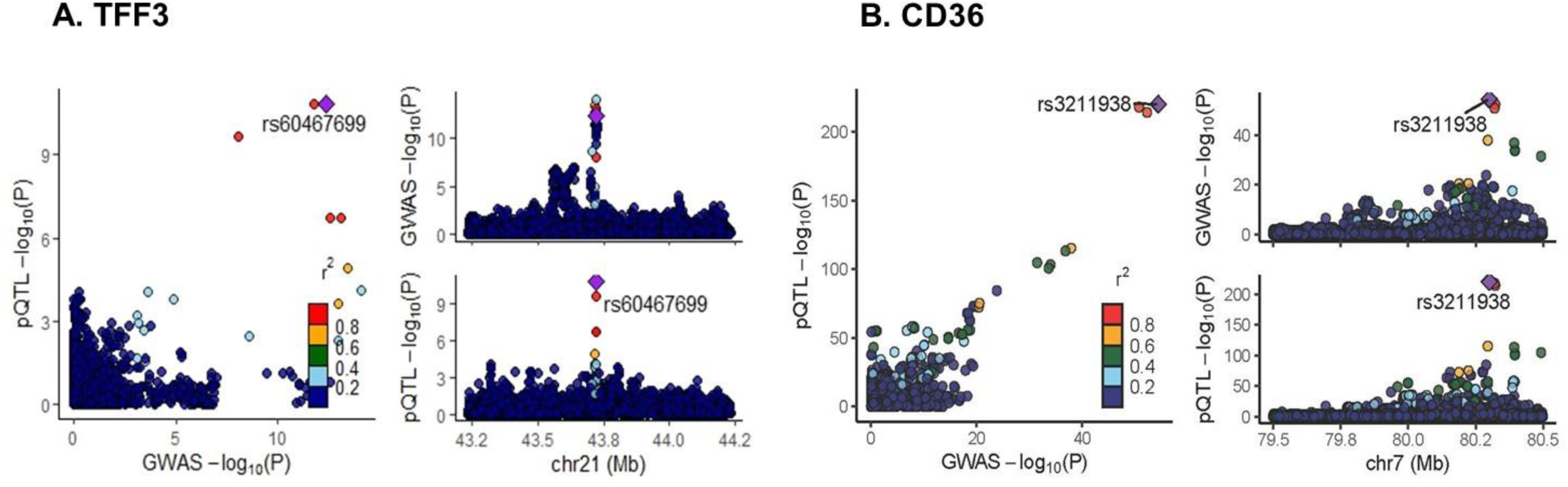
Colocalization analysis of TFF3 (A) and CD36 (B) using meta-analysis of JHS and ARIC pQTL and GWAS of HDL-C. Genetic colocalization analysis of TFF3 and CD36 based on meta-analysis of pQTL results from JHS and ARIC, and GWAS-HDL-C. A. TFF3 rs60467699 significantly colocalized with HDL-C (*P*=4.44E-13) and pQTL (*P*=1.46E-11). B. CD36 rs3211938 significantly colocalized with HDL-C (*P*=4.18E-55) and pQTL (*P*=4.75E-221).

## DISCUSSION

Having previously identified a significant negative association of plasma soluble LAG3 with HDL-C and increased risk for myocardial infarction as well as identifying a LAG3-associated protein network in MESA and FHS^3,16^, the objectives of this current study were: (1) to leverage the datasets available in TOPMed to examine the association of available soluble immune checkpoint proteins by SomaScan (LAG3, CTLA-4, PDCD1LG1, PDCD1LG2, CD40LG, HAVCR2) with each other and their respective associated protein networks with major lipid classes, (2) to examine the association of these immune checkpoint-lipid-associated proteins with all-cause mortality risk, and (3) to identify the overlap of genetic determinants of the mortality associated proteins with genetic variants for HDL-C and LDL-C.

Opdualag is the first FDA approved combination immunotherapy but there are many more clinical trials targeting LAG3 hoping to advance to final approval^1^. We had identified LAG3 and its association with HDL-C based on a previous observation that the *SCARB1* noncoding rs10845677 single nucleotide polymorphism (SNP), which is a long distance enhancer, was an independent factor affecting plasma LAG3 levels^3^. As shown in **Table 3**, a meta-analysis revealed that plasma LAG3, PDCD1LG1, and HAVCR2 were significantly negatively associated with HDL-C. We observed that PDCD1LG1, PDCD1LG2, and HAVCR2 were significantly negatively associated with LDL-C; LAG3, PDCD1LG1, PDCD1LG2, and HAVCR2 were negatively associated with triglycerides, while CD40LG was not associated with any of the lipids. We initially focused our analyses on the LAG3-HDL-C-, PDCD1LG1-HDL-C-, and HAVCR2-HDL-C-associated proteins given our previous findings but also because the mortality risk in patients with elevated HDL-C levels has not been thoroughly investigated, as exemplified by the recent guidelines from the European Society of Cardiology and American professional organizations^34, 35^. A PubMed literature review using search terms “immune checkpoint proteins and cholesterol” identified 156 entries, with some publications focusing on the role of cholesterol in anti-tumor immunity (accessed on November 9, 2023). One such publication reported how the anti-tumor effect of murine CD8^+^ T cells was enhanced by modulating intracellular cholesterol levels^36^. A more clinically relevant publication reviewed the emerging observations of increased CVD events following immune checkpoint blockade, concluding that oncologists should become more vigilant in assessing CVD risk in their patients^37^. Jo et al. recently reviewed the increased association of ICs with atherosclerotic disease, emphasizing the importance of the intersection of lipid metabolism, ICs, and cardiovascular adverse events^38^.

We previously identified a LAG3-associated protein network with HDL-C and mortality risk in MESA and FHS^16^ and have now expanded our observations with inclusion of the JHS and other soluble immune checkpoint proteins. We did observe a direct association of HAVCR2 with all-cause mortality risk. HAVCR2 was first described in 2002 by Monney et al.^39^ as a major regulator of macrophage immune responses. Bailly et al.^40^ has reported that elevated levels of soluble HAVCR2 are associated with chronic autoimmune diseases and viral pathologies but a clear mechanism of action is still unknown. Nonetheless, HAVCR2 (aka TIM3) has advanced as a therapeutic target in at least forty clinical trials (https://clinicaltrials.gov/search?cond=TIM3, accessed September 04, 2024).

When we examined the association of LAG3 (SomaScan) or LAG3-HDL-C associated proteins with mortality risk, we did not observe any significant associations. However, LAG3 measured by ELISA, in the high HDL-C stratified group, did reveal a significant association with mortality risk. The correlation between LAG3 (SomaScan, Exam 1) with LAG3 ELISA (Exam 2) was significant (N=934, beta estimate 0.09, r^2^=0.008, *P*=0.006). We previously reported that plasma LAG3 (ELISA) was positively associated with IL-10 (*P*<0.0001), suggesting greater inflammation in LAG3 deficiency^3^. We have also reported greater CD4^+^ and CD8^+^ T cell infiltration in hypercholesterolemic *Lag3*^-/-^ mice^41^. In mice with Lag3 deficiency in bone marrow-derived dendritic cells, we reported higher glycolytic activity and CD4^+^ T cell proliferation as compared with wild-type mice^42^. We hypothesize that LAG3 deficiency is associated with inflammation and dysfunctional HDL-C leading to increased all-cause mortality risk.

One of the major objectives of this project was to examine overlap of genetic determinants of the mortality associated proteins with genetic signals for HDL-C and LDL-C. As we have shown, in JHS we identified sixty-one HDL-C-mortality proteins and fifty-two LDL-C-mortality proteins. We then performed a meta-analysis using pQTL data from JHS and ARIC to prioritize the genetic colocalization analysis. Of the six HDL-C-mortality associated proteins exhibiting high HDL-C_high mortality risk (REG4, IGFBP1, MIP5, MIC1, TFF3, IGFBP2), the colocalization analysis identified *TFF3* rs60467699 as significantly co-localizing between GWAS-HDL-C and pQTL. There were no PubMed publications linking TFF3 with HDL-C, LAG3, or PDCD1LG1 but TFF3 has been studied in gastrointestinal disorders^43^. We have now identified a novel association of TFF3 with HDL-C consistent with the HDL-C paradox phenotype (higher TFF3 plasma levels with higher HDL-C levels and increased all-cause mortality risk). Of the remaining five proteins, it is possible that causal variants reside outside of the cis window or the circulating levels of these proteins are regulated by post-translational mechanisms. More is known about the *CD36* rs3211938 SNP (common missense T>G SNP with frequency in TOPMed of 0.03%) and its association with inflammatory biomarkers such as C-reactive protein, HDL-C and endothelial dysfunction^44,45–47^. Little is known about the association of CD36 with immune checkpoint proteins *per se* but there are a few publications related to CD36 expression and function in tumor infiltrating CD8^+^ T cells^48,49^. Kummer et al. reported that in a mouse model of Alzheimer’s disease, mice deficient in PD-1 (the receptor for PDCD1LG1) exhibited reduced Cd36 expression on microglial cells^50^. *Cd36* null mice exhibited higher HDL-C levels due to diminished hepatic HDL holoparticle uptake, suggesting this network of ICs with soluble CD36 warrants more mechanistic studies^51^. One publication identified significant co-expression of *Cd36* and *Tff3* in mouse models of hepatic steatosis^52^.

Limitations of the study analysis and results warrants further discussion. In MESA, we have previously published on the plasma LAG3 ELISA assay and the quality metrics for the SomaScan LAG3 measurements^3,16^. Likely due to the larger sample size of plasma LAG3 measured by ELISA, we now observe increased mortality risk in subjects with lower LAG3 and higher HDL-C levels. We had previously identified the *LAG3* rs3782735 variant that was significantly associated with plasma LAG3^16^, and this observation has now been found in JHS. We have focused primarily on associations with HDL-C due to its continued central role in CVD risk assessment, while acknowledging the large body of publications related to other characteristics of HDL, including its size, composition, and cholesterol efflux functionality. The investigation of cholesterol and HDL-C in cancer is receiving traction^53, 54^, and hopefully our identification of possible causal genes associated with immune checkpoint proteins, HDL-C, and mortality risk will spur more mechanistic studies and support from multiple influential partners, including Big Pharma with its growing use of artificial intelligence in drug discovery (https://www.clinicaltrialsarena.com/comment/big-pharma-ai-partnerships/?cf-view)^55^. Non-genetic etiologies could be operative here as well since we observed negative associations of LAG3, PDCD1LG1, and HAVCR2 with total cholesterol, HDL-C and triglycerides, suggestive of an association of these immune checkpoint proteins with hepatic lipase expression and function^56^.

In summary, we have identified novel associations between LAG3, PDCD1LG1, HAVCR2 with HDL-C and mortality risk. We can now include the *TFF3* rs60467699 SNP as a representative of the HDL-C paradox phenotype, with the *CD36* rs3211938 SNP representative of the classic phenotype of low HDL-C and increased mortality risk. With the ultimate goal to improve clinical outcomes by reducing adverse events, identifying biomarker predictors and/or potential therapeutic targets, mechanistic investigations of the interactions between HDL-C and ICs in tumor immunity and vascular responses is warranted.

## Acknowledgements

Molecular data for the Trans-Omics in Precision Medicine (TOPMed) program was supported by the National Heart, Lung and Blood Institute (NHLBI). Core support including centralized genomic read mapping and genotype calling, along with variant quality metrics and filtering were provided by the TOPMed Informatics Research Center (3R01HL-117626-02S1; contract HHSN268201800002I). Core support including phenotype harmonization, data management, sample-identity QC, and general program coordination were provided by the TOPMed Data Coordinating Center (R01HL-120393; U01HL-120393; contract HHSN268201800001I). We gratefully acknowledge the studies and participants who provided biological samples and data for TOPMed.

Support for the Multi-Ethnic Study of Atherosclerosis (MESA) projects are conducted and supported by the National Heart, Lung, and Blood Institute (NHLBI) in collaboration with MESA investigators. Support for MESA is provided by contracts 75N92020D00001, HHSN268201500003I, N01-HC-95159, 75N92020D00005, N01-HC-95160, 75N92020D00002, N01-HC-95161, 75N92020D00003, N01-HC-95162, 75N92020D00006, N01-HC-95163, 75N92020D00004, N01-HC-95164, 75N92020D00007, N01-HC-95165, N01-HC-95166, N01- HC-95167, N01-HC-95168, N01-HC-95169, UL1-TR-000040, UL1-TR-001079, UL1-TR- 001420, UL1TR001881, DK063491, HL148610, and R01HL105756. The authors thank the other investigators, the staff, and the participants of the MESA study for their valuable contributions. A full list of participating MESA investigators and institutes can be found at http://www.mesa-nhlbi.org.

The Jackson Heart Study (JHS) is supported and conducted in collaboration with Jackson State University (HHSN268201800013I), Tougaloo College (HHSN268201800014I), the Mississippi State Department of Health (HHSN268201800015I) and the University of Mississippi Medical Center (HHSN268201800010I, HHSN268201800011I, and HHSN268201800012I) contracts from the National Heart, Lung, and Blood Institute (NHLBI) and the National Institute on Minority Health and Health Disparities (NIMHD). Genome sequencing for JHS (phs000964.v1.p1) was performed at the Northwest Genomics Center (HHSN268201100037C). The authors also wish to thank the staff and participants of the JHS.

The Framingham Heart Study (FHS) acknowledges the support of contracts NO1-HC-25195, HHSN268201500001I and 75N92019D00031 from the National Heart, Lung and Blood Institute and grant supplement R01 HL092577-06S1 for this research. We also acknowledge the dedication of the FHS study participants without whom this research would not be possible.

Genome sequencing for Whole Genome Sequencing and Related Phenotypes in the FHS (phs000974.v1.p1) was performed at the Broad Institute Genomics Platform (3R01HL092577-06S1, 3U54HG003067-12S2).

Dr. Rodriguez was supported by a NIH grant, R21CA279098-01 and the Linda and David Roth Chair for Cardiovascular Research endowment.

